# Altered Amygdala Volumes and Microstructure in Focal Epilepsy Patients with Tonic-Clonic Seizures, Ictal and Post-Ictal Central Apnea

**DOI:** 10.1101/2023.03.16.23287369

**Authors:** Claudia Zeicu, Antoine Legouhy, Catherine A. Scott, Joana F. A. Oliveira, Gavin Winston, John S Duncan, Sjoerd B. Vos, Maria Thom, Samden Lhatoo, Hui Zhang, Ronald M. Harper, Beate Diehl

## Abstract

**Objectives:** Sudden unexpected death in epilepsy (SUDEP) is a leading cause of death for patients with epilepsy; however, the pathophysiology remains unclear. Focal-to-bilateral tonic-clonic seizures (FBTCS) are a major risk factor, and centrally-mediated respiratory depression may increase the risk further. Here, we determined volume and microstructure of the amygdala, a key structure that can trigger apnea in people with focal epilepsy, stratified by presence or absence of FBTCS, ictal central apnea (ICA) and post-ictal central apnea (PICA).

**Methods:** 73 patients with only-focal seizures and 30 with FBTCS recorded during video EEG (VEEG) with respiratory monitoring were recruited prospectively during presurgical investigations. We acquired high-resolution T1-weighted anatomical and multi-shell diffusion images, and computed neurite orientation dispersion and density imaging (NODDI) metrics in all epilepsy patients and 69 healthy controls. Amygdala volumetric and microstructure alterations were compared between healthy subjects, and patients with only-focal seizures or FBTCS The FBTCS group was further subdivided by presence of ICA and PICA, verified by VEEG.

**Results:** Bilateral amygdala volumes were significantly increased in the FBTCS cohort compared to healthy controls and the focal cohort. Patients with recorded PICA had the highest increase in bilateral amygdala volume of the FBTCS cohort.

Amygdala neurite density index (NDI) values were significantly decreased in both the focal and FBTCS groups relative to healthy controls, with values in the FBTCS group being the lowest of the two. The presence of PICA was associated with significantly lower NDI values *vs* the non-apnea FBTCS group (p=0.004).

**Significance:** Individuals with FBTCS and PICA show significantly increased amygdala volumes and disrupted architecture bilaterally, with greater changes on the left side. The structural alterations reflected by NODDI and volume differences may be associated with inappropriate cardiorespiratory patterns mediated by the amygdala, particularly after FBTCS. Determination of amygdala volumetric and architectural changes may assist identification of individuals at risk.

## Introduction

Sudden unexpected death in epilepsy (SUDEP) is a leading cause of premature death in people with epilepsy; however, the pathophysiology behind the fatal events remains unclear (1). The presence of frequent tonic-clonic seizures is a major risk factor (2).

Several mechanisms have been proposed to precipitate SUDEP, including interictal or postictal hypoxemia triggered by apnea, or profound loss of blood pressure elicited by arrhythmia or asystole followed by terminal cardiac arrest (2,3). The incidence and mechanisms of cardiorespiratory arrests in epilepsy monitoring units (MORTEMUS) study (2) highlighted peri-ictal and post-ictal respiratory dysfunction as an initiating abnormality which eventually leads to cardiac arrythmia, asystole and death. A few studies evaluating the incidence of seizures associated with respiratory dysfunction suggested that post-ictal central apnea (PICA) may be a clinical biomarker of SUDEP(3,4).

Patients with epilepsy who succumbed to SUDEP, or those who are in the high-risk category for a fatal outcome, showed significant brain structural alterations in grey matter that serve major roles in maintaining breathing and blood pressure, and in recovery from failure in those systems (5–7). The microstructure of those sites, and how alterations in structure might contribute to ictal central apnea (ICA) or PICA remain to be described.

Temporal lobe epilepsy is the most common form of focal epilepsy, and the hippocampus accounts for the majority of seizure onsets (8). The amygdala, a temporal lobe structure with pronounced hippocampal and other temporal lobe projections, often participates in temporal lobe seizures. Amygdala structures receive widespread projections from additional cortical and subcortical sites. A principal concern in seizures involving the medial temporal structures is the pronounced downstream projections to cardiovascular regulatory sites and respiratory timing and drive areas of the brain stem. Normally, these amygdala influences trigger breathing and cardiovascular responses to affective stimuli, particularly fear and anxiety, but a role in non-emotional breathing and cardiovascular control has been recognized (9– 13).

Stimulation of the human amygdala triggers apnea, sometimes without subject awareness of breathing cessation (14–16), leading to the hypothesis that amygdala influences may affect respiratory recovery following seizures. Those influences may be enhanced or disrupted if the amygdala is damaged by recurrent seizures and epileptogenesis.

The aim was to assess the volume and microstructure alterations of subregions of the amygdala in conjunction with breathing parameters, including ICA and PICA across three groups: healthy controls, participants with focal unaware seizures, and participants with focal to bilateral tonic-clonic seizures (FBTCS). We also examined evidence that inappropriate cardiorespiratory patterns mediated by the amygdala may be used to identify individuals at risk.

## 2. Material and Methods

### 2.1 Study design

Participants were recruited prospectively at the National Hospital for Neurology and Neurosurgery, London, as part of investigation to determine autonomic and imaging biomarkers of SUDEP (Center for SUDEP Research; CSR). All subjects gave written informed consent, and the study was approved by the Research Ethics Committee (19/SW/00071). All enrolled participants with epilepsy had video-EEG (VEEG) and respiratory pattern monitoring using respiratory belts and SpO_2_.

All patients had focal epilepsy and were stratified into those with only focal seizures occurring without generalisation (focal-unaware seizure cohort), and those who had recurrent FBTCS.

Further, included subjects had at least one recorded focal-unaware seizure with adequate respiratory monitoring during video telemetry, or at least one FBTCS. All participants underwent a standardized MRI protocol, and all patients had ongoing seizures at the time of the imaging. Historical seizure type data were evaluated for the full cohort of patients irrespective of seizure type. Patients with lesions on MRI were excluded.

### 2.2. MRI: acquisition and image processing

Images from the study participants were acquired from the same 3T GE MR750 scanner. Participants underwent high-resolution (1mm × 1mm × 1mm) 3D T1-weighted anatomical acquisitions and advanced multi-shell diffusion-weighted imaging (DWI) optimized for NODDI: 11 b=0 and diffusion-weighted images with b=300, 700, 2500 with respectively 8, 32, 64 directions; voxel size: 2mm × 2mm × 2mm.

Amygdala segmentation was performed on the T1-weighted images using Freesurfer (Version 7.0.0, Martinos Center for Biomedical Imaging, Charlestown, MA, USA) (17,18).

Diffusion weighted images were corrected for tissue magnetic susceptibility-induced distortion, subject motion, and eddy current-induced distortions using FSL TOPUP (19) and FSL EDDY (20). The DTI model was fitted through FSL DTIFIT (only using b=0, 300 and 700) from which we extracted mean diffusivity (MD) and fractional anisotropy (FA) maps. The NODDI model was fitted using the NODDI Matlab toolbox (21) to extract orientation dispersion index (ODI) and the neurite density index (NDI) (22). Taking advantage of the FWF computed when fitting NODDI, tissue weighted mean (23) was used as ROI-wise statistic to cope with free water contamination. NODDI aims to provide further information regarding tissue specific indices such as NDI which looks at the density of axons or dendrites and ODI which assesses the extent of axons or dendritic projections being incoherently oriented (non-parallel) (22).

### 2.3. Population

Overall, 154 epilepsy patients completed the full imaging sequencing, and had associated respiratory parameters available. After imaging review, 25 patients were removed due to hippocampal sclerosis. A further 11 patients were excluded from the study due to historical seizure type data (patients with focal-unaware seizures had ongoing FBTCS at the time of imaging). Finally, 8 patients were excluded, as they had no recorded seizure activity on VEEG.

A total of 103 patients with epilepsy and 69 healthy controls were eligible for the study. The cohort was initially divided into 3 groups: healthy controls, focal-unaware seizure cohort (never, or historical FBTCS, but not for several years), and FBTCS cohort with ongoing FBTCS seizures. Subsequently, both focal-unaware seizure cohort and the FBTCS cohort were each divided, depending on the presence or absence of ICA. In a separate analysis, the FBTCS cohort was subdivided depending on the presence or absence of PICA. ICA and PICA presence throughout the cohort was verified by two independent Telemetry Unit neurophysiologists.

Apnea was defined as one or more missed breaths, as in previous studies (4).

### 2.4 Statistical methods

Amygdala volume and microstructure differences between groups were assessed using a multivariate analysis of covariance (MANCOVA), controlling for age and sex. The dependent variables were the diffusion metrics (FA, MD, NDI, ODI) and volume values for each individual participant. The null hypothesis (H0) was that the means across groups were equal for each diffusion metrics and volume. If the MANCOVA values were significant, an analysis of covariance (ANCOVA) was conducted for each diffusion metric or volume individually, controlling for age and sex. Bonferroni correction was used post-test to counteract for multiple comparisons with a family-wise error rate (FWER) of 5%.

The statistical analysis used IBM SPSS Statistics Data Editor (Version 28.0, IBM Corporation, Armonk, NY, USA). The statistical figures were designed using Prism 9 (GraphPad, San Diego, CA, USA).

## 3. Results

### 3.1 Participant characteristics

The participants’ demographics and epileptogenic zones are summarized in Table 1. A one-way ANOVA was performed to compare the participants age across main three cohorts, and showed that the mean ages between at least two groups significantly differed (F (2,170) = [7.584], p = 0.0001). The mean age between groups statistically differed, as demonstrated by an unpaired T-test with Welch’s corrections. NODDI, DTI and volume statistical data used a statistical analysis that corrects for age of the cohort.

**Table 1.** Demographics and epilepsy characteristics.

| **Characteristics** | **FBTCS Seizure Cohort (n=30)** | **Focal Seizure Cohort (n=73)** | **Healthy Controls (n=69)** |
| --- | --- | --- | --- |
| **Age, mean (SD), years** | 30.84 ± 6.38 | 34.58 ± 11.98 | 40.82 ± 12.95 |
| **Sex, n** |  |  |  |
| Male | 22 | 36 | 25 |
| Female | 8 | 38 | 43 |
| **Epileptogenic hemisphere, n (%)** |  |  |  |
| Left hemisphere | 15 (50) | 26 (35.61) | - |
| Right hemisphere | 7 (23.33) | 35 (47.95) | - |
| Multifocal | 5 (16.67) | 10 (13.70) | - |
| Unknown | 3 (10) | 2 (2.74) | - |
| **Epileptogenic zone, n (% of each group)** |  |  |  |
| Temporal onset | 12 (40.01) | 26 (35.61) | - |
| Temporo-occipital | 0 | 2 (2.74) | - |
| Fronto-temporal | 1 (3.33) | 3 (4.11) | - |
| Insula | 0 | 1 (1.37) | - |
| Frontal | 4 (13.33) | 13 (17.81) | - |
| Parieto-occipital | 0 | 1 (1.37) | - |
| Parietal | 1 (3.33) | 0 | - |
| Multifocal | 5 (16.67) | 10 (13.70) | - |
| Hemispheric | 4 (13.33) | 15 (20.55) | - |
| Unknown | 3 (10) | 2 (2.74) | - |
| **Seizures recorded on VEEG, n** | 150 | 451 | - |
| **Participants with ICA, n (% of each group)** | 17 (56.67) | 36 (49.32) | - |
| **Seizures with ICA, n (% of each group)** | 26 (17.33) | 103 (22.84) | - |
| **Duration of ICA, median (range), seconds** | 12.5 (4-38) | 10.5 (4-40) | - |
| **Participants with PICA, n (% of each group)** | 5 (16.67) | - | - |
| **Seizures with PICA, n (% of each group)** | 11 (8.53) | - | - |
| **Duration of PICA, median (range), seconds** | 5 (2-16) | - | - |

Abbreviations: ICA, ictal central apnea; PICA, post-ictal central apnea.

### 3.2 Respiratory data

Data from respiratory bands were available in at least one focal-unaware seizure in the focal seizure cohort, and at least one FBTCS in the FBTCS cohort, overall in 291 of 601 recorded seizures. Pulse oximetry with sufficient quality was recorded in 176 of a total of 601 seizures, from 39 patients. Thirty-six patients with ictal central apnea had ictal oxygen saturation (SpO_2_) available for analysis. Baseline SaO_2_ (M=95.43, SD=1.72) and ICA SaO_2_ (M=88.20, SD=13.90) significantly differed; paired-sample t-test, t(86) = 3.424, p = 0.0009).

### 3.3 Amygdala Volume

Left and right amygdala volumes were initially examined, stratified into three groups: healthy controls, focal-unaware seizure cohort, and FBTCS cohort.

The pairwise comparison post-hoc test with Bonferroni correction showed that the left amygdala volumes were significantly increased in the FBTCS cohort compared to healthy controls (p<0.001) and the focal cohort (p<0.001) (Figure 1. A). Right amygdala volumes in the FBTCS group showed a significant volume increase compared to healthy controls (p<0.001) and the focal-only group (p=0.008) (Figure 1. B). Graphical representations of the mean volume distribution of the left and right amygdala are in Figure 1, while the detailed results are outlined in Supplementary Tables which can be requested from the corresponding author.

**Figure 1.**
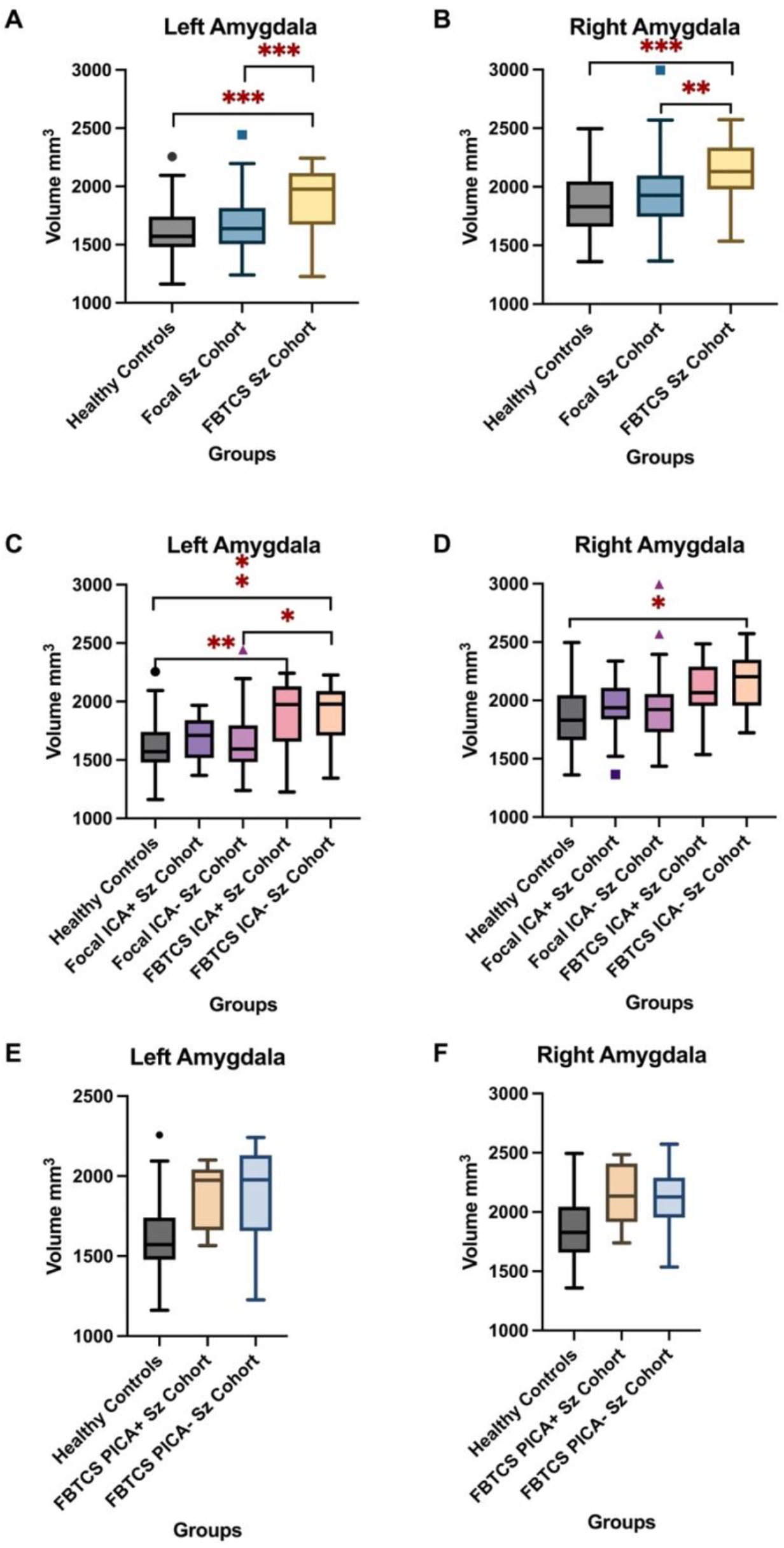
Amygdala Volume Tukey Box Plots. Abbreviations: FBTCS, focal-to-bilateral tonic-clonic seizure; ICA, ictal central apnea; PICA, post-ictal central apnea. (*<0.05, **<0.01, ***<0.001) **(A)** left amygdala volume mm^3^ in 172 participants subdivided into healthy controls, focal-unaware seizure cohort and FBTCS. **(B)** right amygdala volume mm^3^ in 172 participants subdivided into healthy controls, focal-unaware seizure cohort and FBTCS. **(C)** left amygdala volume groups further divided in conjunction with the presence or absence of ICA. **(D)** right amygdala volume groups further divided in conjunction with the presence or absence of ICA. **(E)** left amygdala volume groups further divided in conjunction with the presence or absence of PICA. **(F)** left amygdala volume groups further divided in conjunction with the presence or absence of PICA.

Amygdala volumes were further assessed relative to breathing parameters, and the groups were subdivided according to the presence or absence of ICA. Left amygdala volumes were lowest in the healthy controls, followed by the focal cohort without ICA (Figure 1. C), but not significantly different from healthy controls (p=1.000). The FBTCS cohort without ICA showed a significant volume increase compared to controls (p=0.006), and the focal cohort without ICA (p=0.010). The right amygdala volumes showed a similar distribution, with the FBTCS cohort without ICA showing a significant volume increase compared to healthy controls (p=0.013) (Figure 1. D).

The FBTCS cohort with or without ICA involvement during seizures showed an overall mean volume increase when compared to controls.

The left and right amygdala volumes were examined in association with the PICA identified in the FBTCS cohort (Figure 1. E, F). Comparisons between healthy controls and FBTCS participants were subdivided according to the presence or absence of PICA. In the left amygdala, both the PICA-FBTCS cohort and PICA + FBTCS group showed increased volumes compared to healthy controls (Figure 1. E, F), with only statistically significant volume differences found between healthy controls and the FBTCS group without PICA (p=0.002). Of note, in the right amygdala, both FBTCS groups showed statistically significant volume increases compared to healthy controls.

### 3.4. DTI metrics

DTI showed similar MD and FA values in the left and right amygdala between focal, FBTCS cohorts and healthy controls.

Once the groups were subdivided according to presence of ICA, no statistically significant differences between groups in the left amygdala emerged. However, the right amygdala MD trend was significantly higher in the FBTCS ICA-seizure cohort compared to healthy controls (p=0.014) when adjusted for multiple comparisons using the Bonferroni correction. A similar pattern was found between the focal ICA-cohort and controls (p=0.044).

Split by the presence or absence of PICA, the ANCOVA showed significant differences between groups in the left amygdala in both the MD and FA metrics. The FBTCS PICA+ seizure cohort showed a significantly higher FA than in the FBTCS group (p=0.037). There was no statistical difference between FBTCS PICA+ seizure cohort and healthy controls; however, the FBTCS PICA-seizure cohort had a significantly lower FA when compared to FBTCS PICA+ group (p=0.037).

Significantly lower FA values were found in the FBTCS PICA-seizure cohort when compared to healthy controls (p=0.008). No statistically significant differences emerged between FBTCS PICA+ group and healthy controls in the MD metrics (p=1.000); however, statistically higher MD values appeared in the left amygdala FBTCS PICA-group compared to controls. In the right amygdala, a similar significant increase in MD was found between the FBTCS PICA-group and healthy controls (p=0.022).

### 3.5 NODDI metrics

The left and right amygdala neurite density index (NDI) values were significantly lower in both the focal and FBTCS groups than healthy controls (p<0.001), with the FBTCS group being the lowest of the two (Figure 2. A, B).

**Figure 2.**
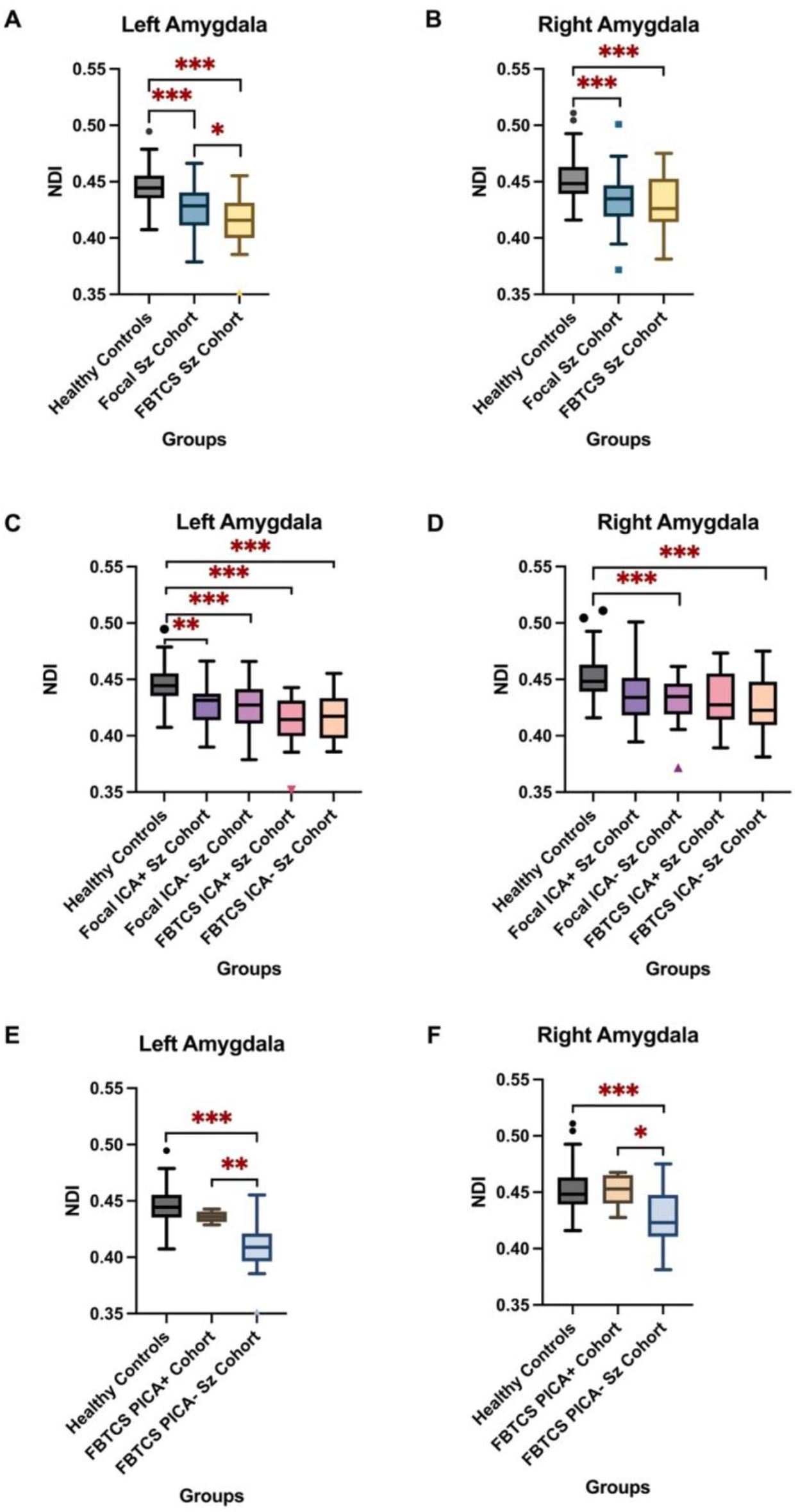
NDI Tukey Box Plots. Abbreviations: NDI, neurite dispersion index; FBTCS, focal to bilateral tonic-clonic seizure; ICA, ictal central apnea; PICA, post-ictal central apnea. (*<0.05, **<0.01, ***<0.001). **(A)** left amygdala NDI in 172 participants subdivided into healthy controls, focal-unaware seizure cohort and FBTCS. **(B)** right amygdala NDI in 172 participants subdivided into healthy controls, focal-unaware seizure cohort and FBTCS. **(C)** left amygdala NDI groups further divided in conjunction with the presence or absence of ICA. **(D)** right amygdala NDI groups further divided in conjunction with the presence or absence of ICA. **(E)** left amygdala NDI groups further divided in conjunction with the presence or absence of PICA. **(F)** left amygdala volume groups further divided in conjunction with the presence or absence of PICA.

A one-way ANCOVA revealed that statistically significant differences in NDI appeared between the four groups and healthy controls once the patient cohort was divided according to ICA (Figure 2. C, D). The most pronounced declines were found between healthy controls *vs* focal ICA-Seizure Cohort (p<0.001), controls *vs* FBTCS ICA+Seizure Cohort (p<0.001), and controls *vs* FBTCS ICA-Seizure Cohort (p<0.001). There were no significant differences between focal ICA+ Seizure Cohort *vs* FBTCS ICA+Seizure Cohort or focal ICA-Seizure Cohort *vs* FBTCS ICA-Seizure Cohort. A significant decline was found in NDI in the PICA subgroup *vs* the TCS group (left amygdala: p=0.004; right amygdala: p=0.042) (Figure 2. E, F).

The orientation dispersion index (ODI) was decreased in both focal and FBTCS groups relative to healthy controls, with the only statistically significant decline found in the left amygdala between the focal seizure cohort and healthy controls (p=0.013). Of note, patients with recorded PICA had the highest decrease in bilateral amygdala ODI values of the population.

## Discussion

### Overview

Nearly a quarter of seizures recorded in this study were accompanied by impaired central breathing control associated with seizure activity, with ICA found in more than half of all cases, a prevalence similar to previous reports (24,25).

The amygdala volume findings show a strong association between the volume of grey matter changes and the presence or absence of FBTCS in conjunction with ICA and PICA, an association not previously reported. A key finding was that patients in the FBTCS PICA group experienced the highest volume gain relative to healthy controls.

Amygdala ODI and NDI were decreased in focal and FBTCS groups relative to healthy controls, most prominently in the FBTCS group.

Detailed demographics of the participants revealed that the mean age statistically differed across the epilepsy groups and the healthy controls. Female-to-male ratios were comparable between the focal-only seizure cohort and healthy controls, with the FBTCS groups comprised of a majority of males. However, as the data analyses were controlled for age and sex, these factors should not contribute to major result differences.

The majority of the focal epilepsy patient cohort had seizure onsets in the temporal lobes; however, a large proportion of the patients with epilepsy had an indeterminate onset which could only be described as hemispheric or multi-focal. Despite the inhomogeneous seizure onset distribution, the amygdala is likely to have been involved throughout the seizure electrical activities *via* the multiple anatomical interactions with other temporal lobe structures.

ICA often accompanies seizure activity in the amygdala (16,26), with direct electrical stimulation inducing apnea (14,16). The findings in functional pathology add to experimental animal evidence that subnuclei within the amygdala can pace inspiratory efforts (27).

### Amygdala Volume and Breathing Disturbances

Significant volume increases in the amygdala appeared in epilepsy patients, with subjects having PICA showing the most extensive increase in volume bilaterally; whereas, patients with only focal seizures had the lowest volume increases. The amygdala volume alterations are relevant, because of its marked role integrating signals provided from a wide range of afferent receptors and projecting signals to lower brain. The central nucleus of the amygdala, for example, receives a wide range of inputs and then projects to the periaqueductal grey and parabrachial pons (28), can influence both overall drive and timing of breathing (27).

Grey matter volume changes also appear within the hippocampus in SUDEP cases compared to low risk participants and healthy controls (5,7). Although the pathological changes related to the volume increases have yet to be described, a variety of processes may underly the changes, including gliosis (29), inflammation causing neuronal architecture disruption (30) or excitotoxic injury (31). The neural processes accompanying the altered volumes may directly influence the amygdala’s influence on both respiratory action and cardiovascular instability as shown here and in (32). Determining the nature of grey matter microstructure changes from tissue samples following surgical resections may further understanding of volume alterations in different epilepsy cohorts.

Patients with epilepsy who respond favourably to antiseizure medications (ASM) have a reduction in temporal lobe volume, including the amygdala, compared to initial brains scans (33). Volume measurements thus may provide insights into the high-risk population for SUDEP, and also may provide valuable information in predicting overall seizure freedom.

### Amygdala Microstructure and Breathing Disturbances

The processes that may underpin the amygdala volume alterations may also be better understood using imaging techniques which can detect neuronal architecture disruption (34) and vascular changes (35).

Diffusion tensor imaging (DTI) showed decreased fractional anisotropy (FA) in the left amygdala, while those FA values were increased on the right, a finding that was more pronounced for the FBTCS cohort with post ictal central apnea. A decline in FA may be caused by Wallerian degeneration (36), CSF contamination or a change in brain tissue organization as a compensatory mechanism (37). Of note, FA may not be able to reliably distinguish dendritic projections and unmyelinated axons (38).

Using NODDI, we were able to disentangle the various factors that may cause an FA reduction noted above. We demonstrated additional microstructure differences across the cohorts in NDI and orientation dispersion index, ODI. The lowest ODI appeared in the focal cohort, an outcome which may be correlated to a reduction in orientation dispersion of the grey matter which may result from reduced dendritic projections (22). The application of NODDI in clinical research has been validated in other conditions (39) showing lower ODI values in multiple sclerosis demyelinating spinal cord histologically (40) and *in vivo* (41).

The ODI results are also associated with decreased NDI which translates to a reduction in neurite density volume particularly found bilaterally in the FBTCS groups. A low NDI may represent severe loss of dendritic and axonal projections, which in turn, can also interfere with ODI values, as the orientation dispersion is correlated with the tissue sampled (41). NODDI offers an excellent opportunity to further understand the neurite architecture in epilepsy without solely relying on pathological studies.

Dendritic projections loss and reduction in orientation dispersion may partially explain some of the ictal and peri-ictal dysfunctions. The slight asymmetry in values, in both volume and microstructure, between the left and right amygdala may pose challenges for autonomic and respiratory control. Contributions to autonomic control are asymmetric in the limbic system; stimulation of the right insula (with direct projections to the right amygdala) leads to hypersympathetic activation (42), while parasympathetic upregulation is largely influenced by the left insula (43). Specifically, the right amygdala grey matter volume increase may contribute to hyper-sympathetic activation, as previous studies observed direct sympathetic upregulation when stimulation is applied to the right insula. One study in particular (44) highlighted that asymmetrical sympathetic activation may contribute to particularly dangerous cardiac arrythmias. However, as opposed to the insula, in the amygdala, the cardiorespiratory consequences do not appear to show laterality (14).

The asymmetry noted in the amygdala volume and microstructure may also result from differential input from other-than-cortical sites, such as the thalamus, parabrachial nucleus (PBN), periaqueductal grey (PAG), and the nucleus of the solitary tract (45). The asymmetrical input may underlie some of the observed differences in laterality of function. While both left and right amygdala are activated to fear responses, the right amygdala appears to play a role in memory retention (46), and the right amygdala is more involved in nociception signaling than the left. Furthermore, an fMRI study examining sex influences on amygdala function revealed more involvement of the left amygdala in arousing memory consolidation in women over men (47). Two impressions arise from these findings. First, the differential laterality of function likely derives from separate inputs to the left *vs* right amygdala, with those influences having the potential to separately modify the extent of injury in an asymmetric fashion to the amygdala, depending on the origin of the influences.

Second, the impact of damage to the left or right amygdala may be expressed differently in behavior or physiological action, depending on laterality of injury.

The amygdala exerts profound influences on the cardiovascular system through projections to structures that regulate blood pressure (48,49). The insula/amygdala control of the baroreflex adjusts heart rate with blood pressure, thus determining cardiac output in response to stressful stimuli (49–51). The amygdala influences are mediated through multiple brainstem structures, including the nucleus of the solitary tract (NTS) and parabrachial pons, and both sympathetic and parasympathetic systems are targets. Projections from the central nucleus of the amygdala to the periaqueductal gray (PAG) and parabrachial pons (52) influence both respiratory drive and patterning, and also modify cardiovascular action (53). The amygdala can influence triggering/not triggering apnea; pulse stimulation of the central nucleus that projects to the parabrachial pons can elicit inspiratory efforts (27), and, through the projections to the periaqueductal gray, support breathing). Those influences place the amygdala volume and microstructure alterations in an environment to influence vital factors that may contribute to increasing the risk of sudden death in epilepsy.

Further studies are required to correlate the changes observed in the amygdala to individual nuclei, particularly the central nucleus, but also subnuclei which influence the central nucleus and can offer more insights concerning cardiac and respiratory regulation.

### Limitations

Since pulse oximetry and respiratory belt signals were not always of adequate quality for many patients undertaking VEEG monitoring, the true prevalence of inappropriate breathing may be higher. The number of participants satisfying criteria for FBTCS associated with post-ictal central apnea was small. Future studies examining seizures and breathing disturbances would likely need to expand the sample size via multi-center collaborations.

### In conclusion

increased bilateral amygdala volumes, accompanied by a decline in ODI and NDI in patients with epilepsy, particularly the FBTCS group, may reflect processes having a direct effect on cardiac and breathing patterns which may increase the risk of SUDEP. The volume and microstructure changes may be mediated by multiple mechanisms, including local inflammation leading to dendritic projections loss or gliosis, or excitotoxicity elicited by excessive external influences to the amygdala. Recognition of amygdala microstructure alterations may assist in identifying individuals with epilepsy who are at a higher risk for SUDEP.

## Data Availability

All data produced in the present study are available upon reasonable request to the authors.

## Acknowledgements

This work has been supported by the National Institute of Neurological Disorders and Stroke, Grant/Award Number: U01-NS090407 (SL, BD, RH, AL), NINDS – NS090405 (SL). GPW was supported by the Medical Research Council (G0802012, MR/M00841X/1). Support was also provided by the National Institute for Health Research and University College London Hospitals Biomedical Research Centre.

The authors acknowledge the facilities and scientific and technical assistance of the National Imaging Facility, a National Collaborative Research Infrastructure Strategy (NCRIS) capability, at the Centre for Microscopy, Characterisation, and Analysis, the University of Western Australia.

## Disclosure of Conflicts of Interest

None of the authors has any conflict of interest to disclose. We confirm that we have read the Journal’s position on issues involved in ethical publication and affirm that this report is consistent with those guidelines.

## Notes

### Competing Interest Statement

The authors have declared no competing interest.

### Author Declarations

South West - Central Bristol Research Ethics Committee of NHS Health Research Authority gave ethical approval for this work. Research Ethics Committee reference 19/SW/00071.

